# “We had to put ourselves in their shoes”: Experiences of Medical Students and ObGyn Residents with a Values Clarification Workshop on Abortion

**DOI:** 10.1101/2023.02.16.23286043

**Authors:** Taryn M. Valley, Elise S. Cowley, Alma Farooque, Zoey B. Shultz, Margaret Williams, Jacquelyn Askins, Amy Godecker, Laura Jacques

**Affiliations:** Department of Obstetrics and Gynecology, School of Medicine and Public Health, University of Wisconsin-Madison, 1010 Mound St., Madison, WI 53715 USA; Department of Anthropology, University of Wisconsin-Madison, 1180 Observatory St., Madison, WI, 53706, USA; Department of Bacteriology, University of Wisconsin-Madison, 1550 Linden Dr., Madison, WI, 53706 USA; Microbiology Doctoral Training Program, University of Wisconsin-Madison, 1550 Linden Dr., Madison, WI, 53706 USA; University of Wisconsin-Madison, School of Medicine and Public Health, 750 Highland Ave, Madison, WI, 53726 USA

## Abstract

**Purpose:** Values clarification workshops on abortion have been shown to increase support for abortion among healthcare workers. However, few studies have examined the impact of values clarification workshops on abortion among medical trainees. This study aimed to understand medical student and obstetrics and gynecology (ObGyn) residents’ experiences with a virtual values clarification workshop on abortion.

**Methods:** Clerkship year medical students and ObGyn residents at four midwestern teaching hospitals were invited to be interviewed about their experiences in a virtual values clarification workshop on abortion from January 2021 through December 2021. A single interviewer conducted interviews via Zoom using a standardized interview guide. Participants were asked to provide feedback and discuss their experiences in the workshop. Four qualitatively trained evaluators coded the interview transcripts in NVivo, using an inductive approach to establish consensus codes then themes.

**Results:** This study interviewed 37 trainees, including 24 medical students and 13 ObGyn residents, as well as five facilitators, between November 2021 and February 2022. Three themes emerged in both trainee groups. First, participants found the workshops *helped trainees clarify and understand their own views on abortion* through thought exploration, peer validation, and reflection on their views’ potential societal impacts. Second, through the workshop, *participants reflected on others’ opinions on abortion* and better understood the spectrum of beliefs their peers held. Finally, participants found the workshops *helped them explore and develop their professional identity as physicians-in-training*, through practicing communication skills and building trust and mutual respect among peers.

**Conclusions:** Medical trainees found values clarification workshops on abortion to be valuable, helping them establish their own beliefs about abortion, contextualize these beliefs among their peers’, and practice professionalism. These findings indicate that values clarification workshops can play a key role in helping medical trainees discuss abortion and prepare for their professional future.

One in four people capable of pregnancy will have an abortion during their lifetime; abortions are one of the most common medical procedures in the United States^1^. Given this ubiquity, abortion education is recommended for medical students and required for Obstetrics and Gynecology (ObGyn) residents^2–4^. However, abortion education is increasingly sparse for both groups of trainees^5,6^. In addition, abortion is highly stigmatized and polarizing, including within healthcare^7–9^.

## Introduction

One intervention shown to improve knowledge about abortion and decrease abortion stigma is values clarification workshops: these sessions allow participants to examine their own values and attitudes and how they are affected by beliefs, ideals, and knowledge^10^. The medical field has utilized values clarification workshops on other stigmatized topics, such as care for patients with HIV, as a way to reduce provider stigma and improve provider attitudes towards patients^11^.

Ipas, an international organization seeking to reduce unsafe abortion, has created a publicly available online toolkit of Values Clarification and Attitude Transformation (VCAT) workshops on abortion^10^. Ipas has shown that these VCAT workshops improved knowledge and attitudes surrounding abortion among international healthcare workers^12^. The World Health Organization recommends VCAT workshops as an essential component of abortion education to decrease stigma and improve abortion access^13^.

Little is known regarding the impact of VCAT workshops on medical trainees in the United States. In particular, no research examines VCAT workshops with medical students or qualitatively assesses their impact. We implemented virtual VCAT workshops on abortion for medical students and ObGyn residents at four midwestern institutions. Subsequently, we sought to understand the impact of these workshops on abortion attitudes and behavioral intentions using qualitative analysis. We interviewed a subset of participants to better understand their experiences with VCAT workshops, and how they perceived the impacts of these workshops on their attitudes, beliefs, and intentions surrounding abortion.

## Methods

### The workshop

From January to December 2021, we conducted a virtual adaptation of the “Four-Corners” VCAT workshop on abortion^10^ at three midwestern medical schools with medical students on their ObGyn clerkships and with ObGyn residents at four midwestern residency programs. Attendance was mandatory for students and residents who were not on vacation or post-call. Prior to the workshop, we emailed two forms, Form A and Form B, to participants. Form A contained 12 statements about personal beliefs about abortion services for others (Supplemental Appendix 1), while Form B contained 12 statements about personal beliefs on abortion services for oneself (Supplemental Appendix 2). Trainees selected the degree to which they agreed or disagreed with these statements on a four-point Likert scale.

Immediately before the workshop began, we sent each participant an anonymous Form A response, which had been filled out by one of their colleagues enrolled in the workshop. We asked trainees to participate in the workshop based on the perspective of the person who had filled out the anonymous Form A they received (not their own views).

The workshop was conducted on Zoom. A trained faculty facilitator greeted the participants and explained the process and purpose of the session. The facilitator then read each of the 12 Form A statements aloud. After each statement was read, the facilitator posted a Zoom poll. Participants responded with their anonymous colleague’s answers from Form A. Participants could see a visual display of the variation in peers’ responses to each question after the polls closed. We selected four statements for small group breakout discussions, where facilitators asked trainees to discuss possible rationales for agreeing or disagreeing with the statements. The workshop concluded with reflection and discussion.

### Interview Participant Selection

In addition to Forms A and B, we invited all workshop participants to complete identical pre-and post-workshop Qualtrics online surveys; participants received ten-dollar Amazon gift cards for each completed survey. These surveys collected demographic information, attitudes toward abortion care, and behavioral intentions for future practice. To assess attitudes toward abortion care, we asked trainees the degree to which they agreed with 17 statements about abortion, using a five-point Likert scale. To assess behavioral intentions, we posed six yes/no questions about trainees’ intent to learn about, advocate for, refer patients for, and provide abortion care^12^. We calculated summative pre-workshop abortion attitude scores ranging from 0 (most negative) to 100 (most positive)^12^. Fifty (70%) ObGyn residents and 307 (57%) medical students completed both the pre-survey and post-surveys.

For medical students, we stratified pre-survey attitude scores by quartile for each of the three institutions. We randomly selected students from the highest and lowest quartiles to participate in a follow-up interview about their experience in the VCAT workshop. Among medical students, we aimed for three completed interviews in each of the first and fourth attitude quartiles, per institution, for a goal of 18 interviews. We also randomly selected students who did not complete the pre or post-surveys (and thus did not have an attitude score) to participate in an interview, in an attempt to mitigate non-response bias. We aimed for three non-responder interviews per institution, for a goal of 27 interviews total (3 students who did not respond to the pre or post survey + 3 students with pre-survey scores in the 1^st^ quartile + 3 students with pre-survey scores in the 4^th^ quartile = 9 interviews * 3 schools = 27 interviews). We used this selection process to achieve representation across institutions, roles (student or resident), as well as the spectrum of abortion attitudes.

For residents, due to overall higher attitude scores and smaller sample size^14^, we stratified based on whether their attitude scores were above or below the median score, within institutions. Due to the limited sample size, we emailed all residents inviting them to be interviewed. For the residents who responded to be interviewed, if they fit into one of our three groups, we selected the first to respond to the invites, with a goal of three completed interviews from each stratum and three non-responder emails per institution for a goal of 36 interviews total (3 residents who did not respond to the pre or post survey + 3 residents with pre-survey scores above median + 3 residents with pre-survey scores below the median = 9 interviews * 4 residency programs = 36 interviews).

We emailed participants from each group in subsequent batches of three until we reached the goal number of interviews or until we sent four rounds of email invites. We selected the first three volunteers of those invited to be interviewed by per participant group (institution, learner level, and attitude score) (Figure 1). Overall, we conducted interviews with 37 trainees: 24 medical students and 13 residents. We also interviewed all five workshop facilitators, but the present study excludes facilitator data. All interview participants, except one facilitator, received a $100 Amazon gift card upon completion of the interview. We conducted interviews from November 2021 to February 2022.

**Figure 1.** Total workshop participants and eventual interviewees for **A**. ObGyn residents and **B**. medical students. Prior to the workshop, all participants were sent a pre-survey to voluntarily fill out. We calculated summative scores on attitudes toward abortion based on the pre-surveys and stratified the attitudes based on quartiles. We invited three groups of participants in both learner groups to be interviewed: 1. all non-responders to the pre- and post-surveys, 2. first (and second in the case of ObGyn residents) quartile attitude scores, and 3. fourth (and third in the case of ObGyn residents) quartile scores. The goal for each group was three interviewees. For ObGyn residents, our goal was 36 interviews total (3 residents who did not respond to the pre or post survey + 3 residents with pre-survey scores above median + 3 residents with pre-survey scores below the median = 9 interviews * 4 residency programs = 36 interviews). While for medical students, our goal was 27 interviews total (3 students who did not respond to the pre or post survey + 3 students with pre-survey scores in the 1^st^ quartile + 3 students with pre-survey scores in the 4^th^ quartile = 9 interviews * 3 schools = 27 interviews). Five facilitators across the three schools were also interviewed, but their interviews were not included for this study. Results are presented as n (%). Pie charts represent the makeup of participants from each program or school for the associated category.

### The Interviews

A single interviewer conducted all the interviews virtually using a standardized interview guide (Supplemental Appendix 3) and audio- or video-recorded the interviews based on participant preference. The interviewer, a departmental communications manager trained and experienced in conducting interviews, was not involved in student or resident education or grading and was blinded to the participants’ attitude scores. The interviewer asked participants to discuss their overall experience in the virtual VCAT workshop, its impact on their own and others’ views and behavioral intentions regarding abortion, and any feedback for improvement. Different members of our team used Zoom auto-transcription and performed confirmatory transcription using ExpressScribe v. 11.10 for Mac (NCH Software, Greenwood Village, CO). We assigned each transcript a unique alphanumeric code and the team member who transcribed the interviews removed extraneous identifying information.

### Qualitative Approach and Analysis

To understand the participant experience with the workshop, we chose qualitative analysis of semi-structured interviews as our methodological approach. Qualitative research is best equipped to provide answers to questions about why, how, and under what circumstances events occur^15^.

We used an inductive qualitative approach, building our understanding from the data. We established a set of codes, then themes, to explore participants’ experiences with VCAT, the virtual format, and the impact the workshop had on their beliefs about abortion. Four qualitatively trained evaluators, not including the interviewer, coded the transcripts in NVivo Release 1.5 (QSR International, Burlington, MA). We coded initial transcripts synchronously to establish consensus and develop a codebook, which the four coders then used to code the remaining transcripts in pairs, ensuring ongoing consensus. At least two team members coded each transcript. Using these codes, we identified salient themes from these codes, first individually, then reached consensus themes as a group. Finally, we identified specific participant quotations that exemplified these themes within their relevant codes. These quotations and broader patterns in the qualitative data make up the results of this qualitative study.

The University of Wisconsin-Madison Health Sciences Institutional Review Board determined this project to be exempt from institutional review (2020-0803).

## Results

Thirty-seven trainees completed interviews, including 13 ObGyn residents (7 with above the median attitude scores, 5 with below the median attitude scores, and one non-responder) and 24 medical students (9 from the 1^st^ quartile of attitude scores, 9 from the 4^th^ quartile, and 6 non-responders) (Figure 1). This sample spanned three medical schools and four residency programs in two states.

We identified three themes present in both medical student and ObGyn resident interviews: participants found that VCAT workshops helped them 1) *clarify their own views on abortion*, 2) *reflect on others’ opinions on abortion*, and 3) *create a professional environment to discuss this often complex and difficult topic*.

### I. VCAT helped participants clarify their own views on abortion – Table 1

Participants shared that the VCAT workshop helped them **clarify and understand their own complex views on abortion**. Participants experienced this clarification of their own views through three main pathways: 1) safe thought exploration, 2) validation by peers, and 3) reflection on the societal impacts of trainees’ views on abortion.

**Table 1:** Subthemes and quotes from VCAT participant interviews supporting Theme 1: *Clarification of personal views about abortion*.

| Subtheme | Quote Number | Quote |
| --- | --- | --- |
| VCAT as a safe opportunity for thought exploration | T1.1 | “I think that my views of abortion have changed... from a Christian background, my views on abortion coming in were essentially, in most cases, I think we should avoid abortion, if possible. Maybe that statement I don't even disagree with now, but I just recognize that I'm wrong about so much stuff in life, and there's just so little that I know about life except for the people I'm very close with in life, like what they're truly going through and where they're coming from... I just have a lot of respect for other viewpoints and I just see a lot of hypocrisy ... if I'm going to view abortion that way, then there's a whole train of things that come unraveled...so I think that tightness with which I grip my viewpoints and want to put them on other people very much loosened.” (M2-01) |
|  | T1.2 | “I was maybe a little bit more neutral coming in, as opposed to pro-life, pro-choice and you know, after hearing a lot of my classmates arguments or other people's arguments towards the pro-choice, it really just made a lot of sense to me, like I don't know why I've never gone this deep into thinking about it... after kind of going through that, and then hearing my more neutral responses, like I said it was kind of eye-opening and I kind of wonder... why did I answer that more neutrally when like clearly I am... more towards the pro-choice and women having more say over the process than they do.” (M2-09) |
|  | T1.3 | “[Participating in values clarification workshops] always makes me realize how much the idea of choice is a lot more nuanced than it can seem from like the political perspective... every time I do values clarification, I remember there's such a broad spectrum of values within, even within people who are purportedly pro-choice. And it always makes me think more hard [sic] and more intentionally about, where do I lie in that nuanced spectrum... it's always really enlightening to sort of have to interrogate that for myself through things like this.” (M2-03) |
|  | T1.4 | “[E]ach time I've sat down to do this activity... I articulate to myself... stronger and more nuanced and firmer beliefs in my personal comfort level, and my personal dedication to abortion care... in earlier versions, pre-residency versions of a values clarification exercise, I think I theoretically agreed with the, at least what I interpret to be widely held belief that abortion becomes more controversial or less palatable or harder to stomach... at more and more advanced gestational ages... |
|  |  | through this workshop and through kind of other reflection experiences [I've] more recently come to understand... I am comfortable with this care being provided at essentially any gestational age for essentially any indication, because I have seen so many examples that I don't think someone who's having some theoretical non-informed conversation about this type of care could even fathom.” (R2-03) |
| Clarification of personal abortion views through peer validation | T1.5 | “[N]ever once during the session did I feel like it was confrontational or judgmental... [the person] that I was representing was someone that had similar thoughts and beliefs to my own so being able to articulate and represent them wasn't that far off from what I already thought... I've never done a values clarification before, but in other sort of group discussions about topics like this that have very polarizing sides, I feel like it becomes more of a debate than it does like an exploration of thought.” (R1-04) |
|  | T1.6 | “When other people are representing my beliefs... people that were representing that point offered many of the same arguments I would offer, and there was [sic] also times where a lens I didn't realize was used in support of it, which was really cool to hear.” (M3-01) |
|  | T1.7 | “I tend to see the gray... in the argument... [VCAT] showed to me that other people feel that way too, which is kind of nice... sometimes because I didn't feel like I was solidly in like a pro-life or a pro-choice camp that you know, I was kind of alone in that, and seeing that other people in some ways feel that way too was kind of nice.” (M1-07) |
| Consideration of the societal impacts of personal abortion views | T1.8 | “[I]t seems in talking with classmates... after this workshop that amongst medical students, that there is a little bit more acceptance, maybe than before, and trying to destigmatize abortion and access to abortion... I'm pretty sure the majority of the United States is in favor of access to legal abortion... And I think these conversations help destigmatize that.” (M2-06) |
|  | T1.9 | “[F]or most people a lot of these beliefs are pretty rooted to cultural or background or religious contexts, which won't, which often don't change... while their opinion of it might not change, I think if they have more information about it, it's less likely to be stigmatized... even if you can't change someone's mind, maybe you can change the public's perception by just providing information, so I think sessions like this are pretty important for that.” (M3-01) |
|  | T1.10 | “I think we have huge power, one way or another. I think that any provider can, whether or not they're in ObGyn, can experience patients who are in difficult situations or who are in unplanned or unwanted pregnancies, and our language and how we talk with them and what services we offer them and how we frame that language and that discussion can really impact that one individual, but |
|  |  | also you know the greater society, because if that one individual thinks we want to normalize abortion or anti-abortion, it kind of has a rippling effect.” (M3-06) |
|  | T1.11 | “[VCAT has] motivated me to be more involved, for instance, like when we've had opportunities [for advocacy]. So, in terms of looking at abortion care as kind of, patient doctor relationship and focusing on compassion and empathy in those patient interactions as well as looking at the societal factors that also play into that, and being more motivated to advocate for better access to care for patients. I think it's mostly just re-incentivized the importance of those things in the different spheres.” (R1-02) |
|  | T1.12 | “[VCAT] energizes you to be more proactive about like the legality of things and how like our jobs and our livelihood is like very much affected by the legislature and that's like a very threatening thing...the abortion training workshop just kind of reminds you of this, I don't think that we necessarily discussed it, but I think...this is something that I do believe, and I should be more proactive and you know, taking a stance.” (R2-05) |
|  | T1.13 | “[I]t's good for me to think about this... if I am training to be an ObGyn if I opt out of learning how to do it... I don't think I would be necessarily the best ObGyn [I] can be... I guess maybe this is why the responsibility question kind of really did resonate with me, because I do feel like it is important ... in today's society... If there are ObGyns or doctors who don't train how to do it, or don't... want to learn more about abortion care, I do kind of feel like it just adds to the stigma. So I feel like the session was important in just kind of bringing that to light.” (R3-01) |

The code in parentheses following each quote identifies the speaker as either a medical student (M) or resident (R) from institution 1, 2, 3, or 4 followed by a number assigned to that individual. For example, R2-05 is the 5<sup>th</sup> resident interviewee from institution 2.

First, participants described VCAT as a novel opportunity for **safe thought exploration**. Some people described that their feelings on abortion changed due to their experience in the workshop. For one medical student, through the workshop, “that tightness with which I grip my viewpoints and want to put them on other people very much loosened.” This loosening resulted in a shift in their perspective on abortion (T1.1). Another student shared that their perspective on abortion changed after the workshop, becoming firmer and less neutral than their prior ideas (T1.2).

Overall, though, few participants described their own views as having *changed* across the VCAT experience. Instead, most participants explained that they deepened their *understanding* of their own views, by exploring them through the unique VCAT format. One student, who had previous experience with values clarification exercises, said that going through another VCAT workshop made them more comfortable with the nuance and “broad spectrum” of values about abortion (T1.3). A resident who also had prior experience with values clarification exercises spoke specifically about their expanding comfort with abortion provision across gestational ages, after exploring their own values in VCAT (T1.4). These trainees, whether they felt their opinions on abortion had shifted or not, shared that the VCAT format allowed them to reflect on their own opinions about abortion; participants found this exploration of their own thoughts valuable.

In addition to reflection, the second way participants clarified their own views was through **peer validation** built into the VCAT format. One resident found the session structure supported sharing similar opinions with co-residents: “[N]ever once during the session did I feel like it was confrontational or judgmental” (T1.5). Multiple students and residents shared how they learned from hearing colleagues represent their own opinions. One student enjoyed hearing their own values represented through a different “lens” (T1.6). Many interviewees explained that colleagues validated their specific opinions around abortion as a result of the VCAT format, for example, one student appreciated that other students also felt they didn’t fit strongly in either “camp” regarding abortion (T1.7).

Finally, VCAT made trainees further consider the **societal impacts** of their opinions and communication about abortion care. For example, many participants pointed out that the VCAT workshop itself normalized and destigmatized abortion as a part of routine health care (T1.8-9). Considering this larger scale allowed participants to clarify their own beliefs in the context of complex cultural dynamics surrounding abortion. One medical student reflected that overall, healthcare providers “have huge power” amidst “the greater society” around abortion issues (T1.10). A resident shared how the VCAT workshop changed their perspective on their role in larger societal processes related to abortion, beyond patient care: they said VCAT “motivated me to be more involved, for instance, like when we’ve had opportunities…to advocate for better access to care for patients” (T1.11). Interviewees agreed that VCAT made them feel more invested in abortion topics overall. For example, another resident shared that VCAT encouraged them to be “more proactive” in “taking a stance” regarding abortion (T1.12). Finally, a resident reflected that VCAT helped them think about how trainee choices can impact broader abortion trends and stigma (T1.13).

Overall, medical students and residents shared with interviewers that they better understood their own opinions after the VCAT workshop, including how those opinions were shared by colleagues and how their opinions bear weight in society.

### II. Reflect on others’ opinions – Table 2

Participants described three ways in which the VCAT workshop helped them **reflect on the opinion of others:**1) hearing peer attitudes towards abortion care, 2) benefiting from cognitive flexibility via the VCAT format, 3) speaking freely by virtue of the anonymous structure of VCAT, and 3) finding comfort in shared views.

**Table 2.** Subthemes and quotes from VCAT participant interviews supporting Theme 2: *Reflection on others’ opinions about abortion*.

| Subtheme | Quote Number | Quote |
| --- | --- | --- |
| Reflect on peers' attitudes toward abortion | T2.1 | "[B]efore the workshop, I thought that it would be a little more split, pro- versus anti-abortion, and [in] the workshop... more people were pro-abortion than I would have thought. But people didn't hold as strong of beliefs as I thought. There was, you know, strongly disagree, disagree, neutral, agree, and strongly agree. And a lot of people put agree and disagree, versus strongly agree and strongly disagree, so that also surprised me." (M3-06) |
|  | T2.2 | "Not everybody has very strong views on abortion...I always see it as one camp or the other...but I guess that there's definitely a lot more gray area in there." (M1-03) |
|  | T2.3 | "[W]e'd go around the room, sometimes it would just kind of sound like we all were reading off the same piece of paper... I don't remember somebody like representing my views because if they did... I didn't even recognize that they were mine. But I knew they had to be in there, somewhere, because I was one of the two people that disagreed with the group." (M1-06) |
| VCAT's structure enabled cognitive flexibility, allowing participants to take the view of a colleague | T2.4 | "We had to kind of put ourselves in their shoes and explain the responses of one of our classmates that may or may not have a similar viewpoint to our own." (M1-06) |
|  | T2.5 | "Having a stance that doesn't necessarily have to be your own makes it a lot easier, because then you're not necessarily defending or attacking the student, you're defending or attacking their points, which is a really important part of arguing things like this, because if you're representing someone else's, even if they do happen to coincide with your beliefs, you can kind of detach yourself from that stance." (M3-01) |
|  | T2.6 | "[T]he workshop started with, we all have different beliefs...we were talking about the beliefs of someone else rather than our own with everything being kind of anonymous, so it allowed us to just kind of delve into the topic without getting up in arms or attacking people's personal views." (M3-06) |
|  | T2.7 | "[I]t's obviously a very loaded topic... having the opportunity to be told to represent an opinion that maybe isn't entirely what you believe, but there might be parts of it that you think are true... sometimes I think that those parts of the contrary opinion that I agree with somewhat might not be |
|  |  | accepted by everybody in a group... being in a position to... defend that opinion is really helpful. Because you can make your best argument for it and hear what people have to say about it, without having the other people in the group necessarily tie that opinion to you.” (R1-01) |
|  | T2.8 | “[T]he opportunity to talk about things that are very highly charged in an anonymous way... giving the answers of... co-residents as opposed to my own answers, and seeing how our class feels about these different things, and then also having the opportunity to maybe defend opinions that I don't personally hold, was really helpful to get me to think outside of the box ... [there are] a lot of people that agree about certain things around abortion and to kind of consider possible other ideas was useful in strengthening my own opinions.” (R2-01) |
| VCAT's anonymous structure helped neutralize difficult conversations and allowed participants to discuss topics more freely | T2.9 | “[The] anonymous part I think helped a lot... sharing those [views] can be intimidating for people so being able to submit them anonymously actually revealed the true diversity in our own thoughts, which I think was really good for people to recognize. Sometimes it's easy to think that everybody else thinks like you... I gained respect for a lot of my classmates in terms of knowing where they stood on the for or against abortion line and then seeing them so willingly engage in the conversation... to see the actual diversity of that truly represented, and then to discuss that... it was enlightening for me. (M1-07) |
|  | T2.10 | ” I was a little bit embarrassed about my lack of research beforehand or my lack of knowledge, I was a little bit glad that my initial responses were anonymous... I thought going in you'd be arguing your own points, but you know, being able to argue somebody else's and having the preface of like, we know these aren't your answers, but just do your best to like argue their point and like try to take this side. It was definitely interesting to kind of see that and then not be able to label the person arguing it with that mindset. A And so it kind of provided clear, well as clear as possible, arguments for both sides, and then the person doing it didn't have to feel like ashamed or embarrassed of what they were doing.” (M2-09) |
|  | T2.11 | “[W]e were given the views ahead of time... what we're told to say, and I liked that, because it was anonymous, and you had people defending the views in a way where it felt a little bit more safe rather than kind of just feeling like you were attacked for believing one way or another. And so I did come away thinking it was it was a more safe environment to talk about such a topic.” (M2-06) |
|  | T2.12 | “And so it was nice to... acknowledge some of those things and talk about that was helpful, but in a way, where it didn't like force people to call on people to share about what their opinion is and articulate that... that can be [a] challenging and potentially uncomfortable and volatile situation... so this was a way of having a conversation around this that felt comfortable and safe and didn't feel like anybody was being called out for holding one belief versus another not... [or] far on one side or |
|  |  | another. So I felt like it was it was engaging, it was interesting, but it was also like a safe space to have the conversation.” (M3-05) |
| Comfort in shared views among co-residents | T2.13 | “It was nice to know that kind of the pervasive viewpoint and the vocal majority of my residency program was very pro-choice and very comfortable with the idea of learning about providing abortion care. That was really important to me in a residency program as someone who wants to be an abortion provider... it was hard to tell in the residency interview process like how people really felt... a program provided comprehensive contraception and abortion training, in theory, but it was hard to really know what the culture among the residents was... Going through the workshop, perhaps more than being a huge sort of new exploration of my own values, was a pleasant surprise and good confirmation that the people around me in my residency showed those values.” (R2-03) |
|  | T2.14 | “I think there's a sense of community after participating in a workshop like this... you kind of think you know how other people feel about it [abortion]. But when there's not a space to discuss it, you don't necessarily know how your other co-residents are going to feel. So knowing that there are so many other co residents in my like immediate circle that feel that this is an important service to provide makes me feel confident, if I go into fellowship and don't have the chance to provide abortions, based on what my training ends up being, that there are so many colleagues that I can refer them to.” (R2-01) |
|  | T2.15 | “There were several responses that made it seem like people were comfortable, or not comfortable, but people were providing abortions, because they knew it was something that that they needed to do, but they acknowledge that it made them uncomfortable or they acknowledge that they were... morally grappling with it, despite it being a procedure that they still participated in and learned about. So that was interesting.” (R1-04) |
|  | T2.16 | “[VCAT] was just a reminder that you know destigmatizing, it normalizing it, having these conversations, recognizing that you can feel a little bit ambivalent about the morality of it and still provide this service, I think that was really helpful from this... workshop for counseling patients in the future.” (R2-01) |

Participating in the VCAT workshop allowed trainees to **hear about their peers’ attitudes towards abortion care**. Many participants discussed feeling surprised by the lack of polarity and the broader spectrum of beliefs that the workshop elicited: “people didn’t hold as strong of beliefs as I thought,” said one student, while another expressed surprise about participants’ nuanced views when they expected all participants would fall into “one camp or the other” (T2.1-2). However, some trainees felt that group discussions were limited by dominant or homogenous opinions (T2.3).

Participants described benefit from the **cognitive flexibility component of VCAT**, where trainees participate in the workshop from the standpoint of a classmate. They found it helpful to distance themselves from their own opinions by representing others’ views. One participant described the experience as putting themselves in others’ shoes (T2.4); several other participants echoed this sentiment. The format eased the typically charged and polarizing nature of abortion discussions; many interviewees commented that this shift in tone enhanced the discussion. This cognitive flexibility aspect, taking on the perspective of someone else in their group, allowed trainees to understand others’ views on abortion more deeply: “We were talking about the beliefs of someone else rather than our own,” which “allowed us to just kind of delve into the topic without getting up in arms or attacking people’s personal views” (T2.5-7). Interviewees also explained how this perspective-taking built into VCAT’s format helped them “think outside of the box” about others’ feelings; therefore, trainees’ understandings of other views once again helped them better understand their own views (T2.8).

While trainees experimented with others’ views, the **anonymity of the workshop** allowed participants to speak more freely. Participants described the anonymity involved with the format of the workshop as making the conversation on abortion more neutral. One participant found the anonymous nature of VCAT “enlightening”, helping to reveal the “true diversity” in the group’s opinions on abortion (T2.9). For interviewees, VCAT’s anonymity removed shame and embarrassment from the conversation and helped participants avoid personalized judgements as they deepened their understanding of others’ views (T2.10). Participants cited this “safe environment for discussion” as one reason they and their peers could engage enthusiastically in the workshop (T2.11-12).

Considering differences in trainee levels, medical students were more likely to consider the above workshop dynamics and *how* their peers expressed differing views, residents were more focused on *what* views their peers held. Residents appreciated the opportunity to discuss various attitudes on abortion with their co-residents. Many felt **comforted, validated, and inspired** by their colleagues’ passion about access to abortion services. For instance, one resident expressed “pleasant surprise” that “the vocal majority” of their peers were “very comfortable with the idea of learning about providing abortion care” (T2.13). Residents also reflected on their own decisions, and the decisions of their peers, about whether to provide abortion care in their future careers. Residents who did not plan to provide abortions, but supported abortion access, expressed feeling reassured to hear that many of their colleagues intended to offer abortion care in their future practice (T2.14). Other residents reflected that although some of their peers expressed discomfort or moral ambiguity about abortion provision, residents agreed overall on the importance of abortion provision (T2.15-16).

Overall, participants in the VCAT workshop expressed that VCAT’s anonymous format allowed them to explore others’ perspectives in unique ways. Medical students tended to reflect on workshop dynamics and structure, while residents commented on the implications of their co-residents’ perspectives on future practice.

### III. VCAT helped participants’ professional development – Table 3

The VCAT workshop helped trainees’ **professional development**. They discussed: 1) deepening professional identity and behavior, 2) practicing professional communication skills, 3) sharing values to build trust and mutual respect, and 4) reinforcing the importance of abortion education in professional development.

**Table 3.**
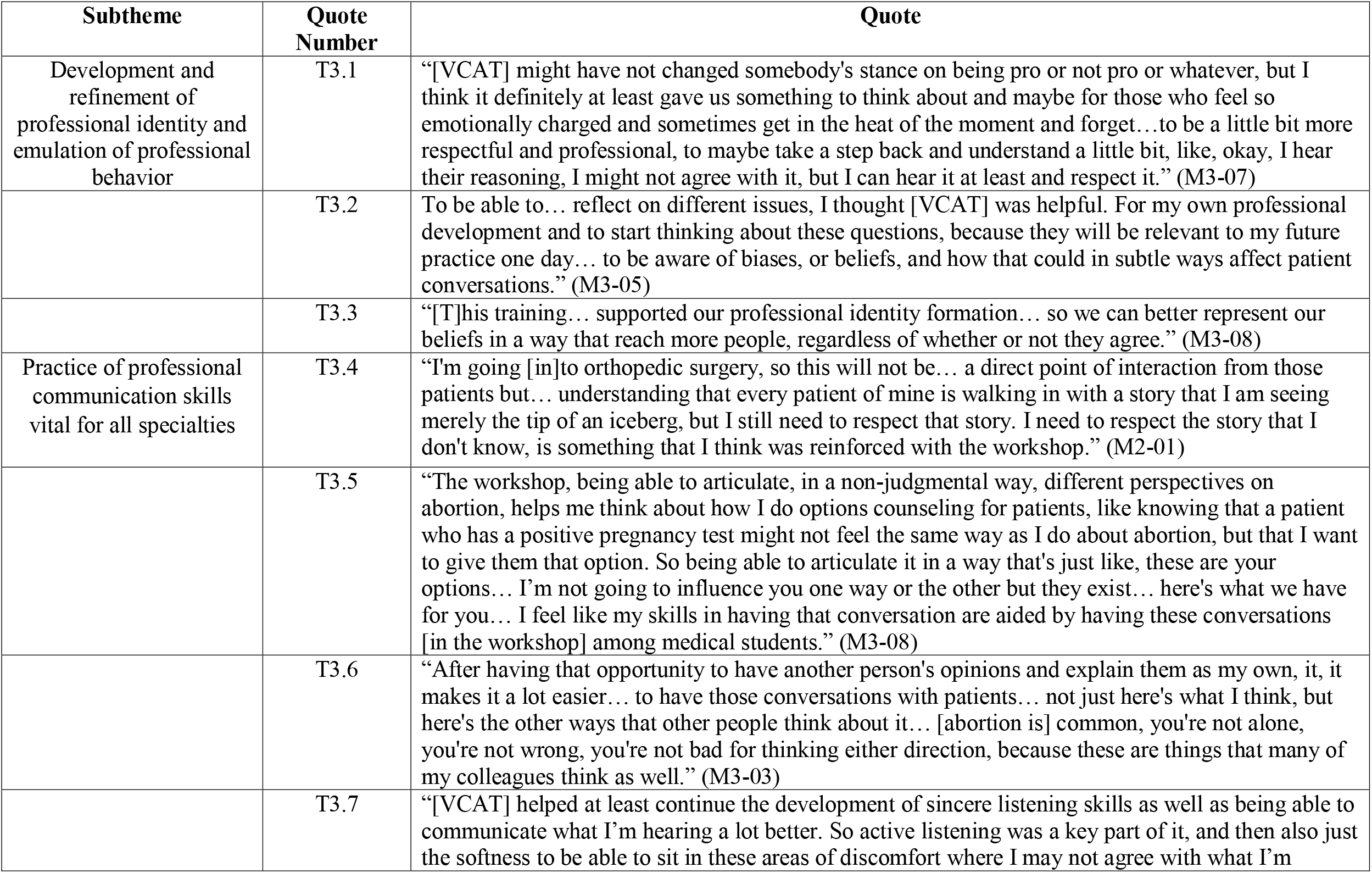
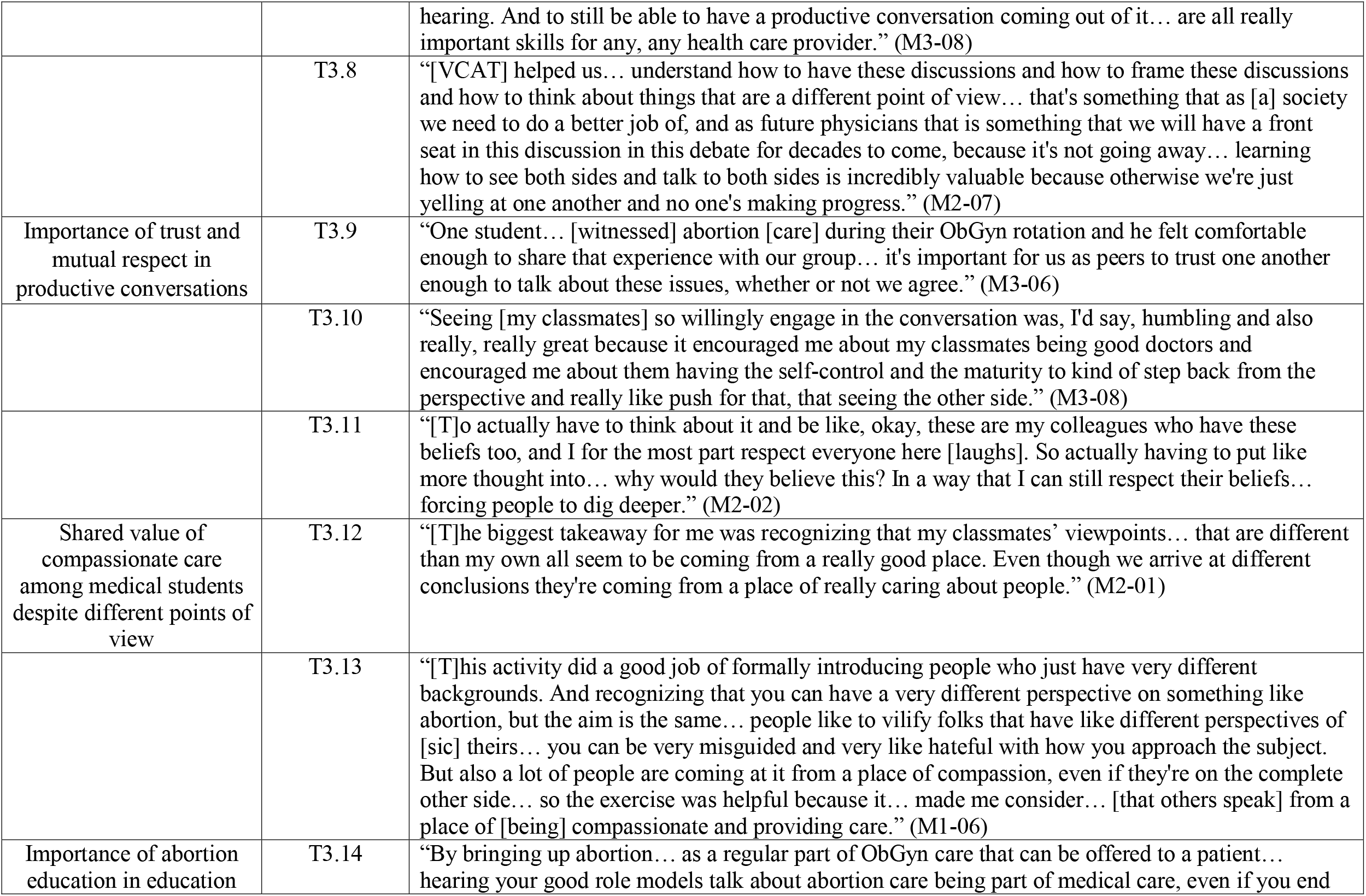
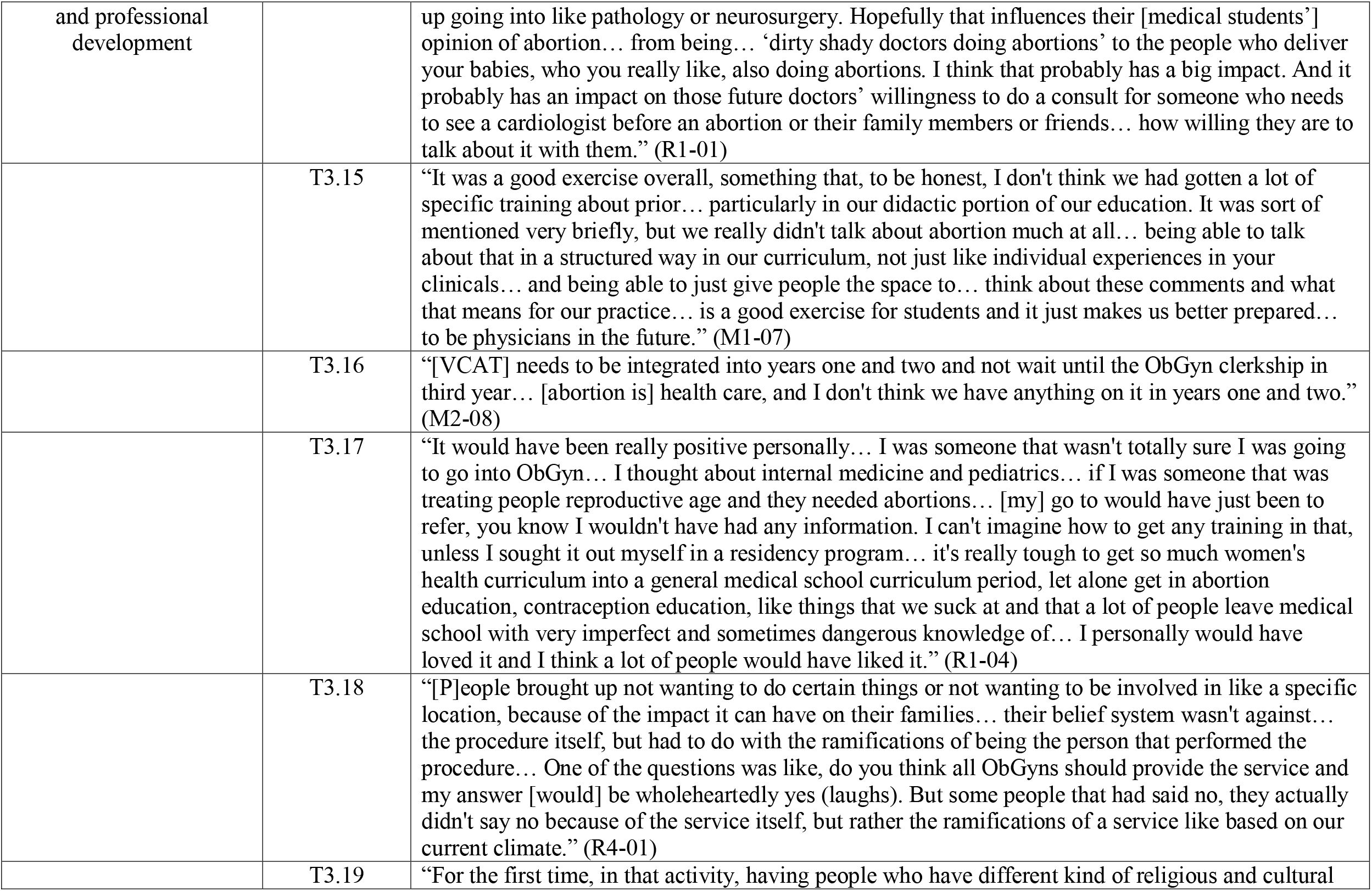
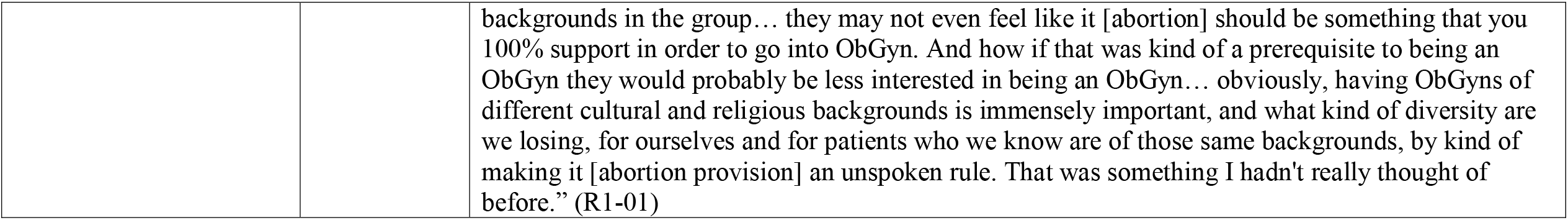
Subthemes and quotes from VCAT participant interviews supporting Theme 3: *Professional Development*.

Participants found that the structure of VCAT allowed trainees to behave professionally around topics that are normally “charged” (T3.1). The interviewees commented on the various ways in which the workshop supported the development of their **professional identity** by discussing questions “relevant to my future practice one day” (T3.2-3). Specifically, many participants found that the workshop prompted them to practice vital **professional communication skills** that they will utilize throughout their career, regardless of specialty (T3.4). One student saw VCAT help students practice “active listening” as well as developing “the softness to be able to sit in these areas of discomfort where I may not agree with what I’m hearing. And to still be able to have a productive conversation coming out of it are all really important skills” in professional development for “any health care provider.” Multiple participants noted that practicing these communication skills through VCAT would improve their patient communication, care, and counseling in the future (T3.5-6). Interviewees also discussed how participants in the workshop practiced navigating disagreement, leaving participants more equipped to disagree respectfully, in and beyond patient care (T3.7-8).

Medical students consistently commented on their peers’ high levels of engagement and comfort sharing personal experiences during the workshop. By sharing their values, they practiced **trust and mutual respect** in productive conversations (T3.9). These dynamics allowed participants who held less-common opinions to feel comfortable and respected; one student said that watching classmates “willingly engage” in respectful discussion “encouraged me about my classmates being good doctors” with “the self-control and the maturity to kind of step back” during charged discussions (T3.10). Another noted that students may have thought through opposing viewpoints with a more nuanced lens because they hold their classmates in high esteem (T3.11). Medical students also reflected on how these respectful discussions highlighted participants’ shared value of compassionate care, despite differing beliefs about abortion: one student realized that “a lot of people are coming at” conversations about abortion “from a place of compassion, even if they’re on the complete other side” of the political spectrum (T3.12-13).

Many interviewees also mentioned **abortion education in relation to professional development**. One resident thought VCAT workshops for medical students would foster future collaboration for people seeking abortion care (T3.14). In reflecting on their professional journeys, medical students often commented on their lack of exposure to abortion care during medical school. Many said that they wanted medical school curricula to include abortion, including one medical student who advocated for incorporating abortion content “in a structured way in our curriculum” beyond individual clinical experiences. They believed formal content like VCAT “is a good exercise for students and it just makes us better prepared” for their careers (T3.15). Another medical student wanted the VCAT workshop to occur earlier during medical school prior to clerkships (T3.16). A resident reflected that, despite the challenges of adding abortion content to curricula, VCAT could have enriched their education when they were a medical student (T3.17). Other residents spoke about new insights they gained during the workshop regarding providing abortion services, which many believed should be an inherent part of the professional identity of ObGyns. Some residents were surprised by the gap they noticed in the workshop between their co-residents’ theoretical beliefs in the right to abortion access, and with their practical plans regarding providing abortion themselves. One resident reflected that some peers did not think all ObGyns should be required to provide abortion services, but not “because of the service itself, but rather the ramifications of a service based on our current climate” around abortion (T3.18). Another resident reflected on the importance of diversity in their profession, with physicians from cultures holding different ideas about abortion; this caused them to reconsider their previous belief that abortion provision should be required for practicing ObGyns (T3.19).

Discussing the nuances of professional identity in the VCAT workshop helped residents understand their own and their peers’ priorities around abortion care. Overall, trainees found that VCAT helped them explore their role as physicians in relation to abortion care.

## Discussion

We found that medical students and residents valued the VCAT workshop experience, as it allowed them to better define their own values towards abortion, understand differing opinions about abortion, and practice professional conversation skills.

There is one published study assessing the impact of abortion VCAT workshops on medical trainees in the U.S^16^. Researchers conducted a VCAT on abortion with ObGyn residents at religiously affiliated institutions and then analyzed the impact by comparing pre- and post-workshop surveys on attitudes towards abortion. Post-surveys demonstrated more supportive attitudes toward abortion after participating in a VCAT workshop^16^. The only qualitative analysis of VCAT workshops on abortion included 20 interviews conducted with healthcare providers in Pakistan after they completed a VCAT workshop on abortion^17^. Providers there reported an increase in professional responsibility to refer for and provide abortion care. They also described feeling more empathy towards their patients because of the workshop^17^. Our study adds to the existing literature by providing additional evidence that participating in VCAT workshops leads to more supportive attitudes towards abortion. Additionally, our qualitative analysis gives insight into *why* attitudes change. Medical trainees we interviewed saw the workshop as a rare opportunity to discuss abortion in a safe space. Participants felt that VCAT’s unique format, particularly its anonymity, helped trainees explore ideas without feeling stigmatized or judgmental themselves.

Strengths of our study include its multi-institutional design, inclusion of both medical students and residents, interviewees across the abortion attitude spectrum, and rich thematic saturation. Limitations of our study include potential responder bias, in that our study did not reach our intended number of participants for 9/21 (43%) of the interview groups, including the majority of the resident groups and the medical student non-responder groups.

Abortion stigma is only likely only to worsen after the recent Supreme Court decision in *Dobbs vs Jackson Women’s Health Organization*. Our findings demonstrate that the values clarification workshop structure can allow trainees to engage with difficult, stigmatized topics. The skills that trainees we interviewed described—communicating respectfully, embracing nuance, and putting themselves in others’ shoes—are crucial to physician abortion training but are not limited to this sphere. This workshop could be adapted to instruct any number of content areas, such as addiction, mental healthcare, and gun violence, among others. Further research could explore the use of the VCAT format for other stigmatized topics in medicine as well as for other types of healthcare providers.

## Supporting information

Supplemental_Appendix_2_FourCorners_FormB

Supplemental_Appendix_1_FourCorners_FormA

Supplemental_Appendix_3_FINAL_VCAT_InterviewGuide

## Data Availability

All data produced in the present study are available upon reasonable request to the corresponding author.

## Acknowledgements and Disclosures

The authors thank Andrea Zorbas, Sharon Blohowiak, Amanda Wildenberg, and Kelly Winum for administrative support. The authors also thank Nathan Jones and the UW-Madison Survey Center.

This research did not receive any specific grant from funding agencies in the public, commercial, or not-for-profit sectors. TMV is a Medical Scientist Training Program (MSTP) student funded in part by the CORE Lab, funded by a large, anonymous family foundation, and in part by Medical Scientist Training Program grant T32GM140935. ESC is an MSTP student and was supported by an NLM training grant to the Computation and Informatics in Biology and Medicine Training Program (NLM 5T15LM007359) at UW-Madison, and in part by MSTP grant T32GM140935.

None of the authors have a financial or other conflict of interest.

Ethical approval has been waived for this study by the University of Wisconsin-Madison

Minimal Risk IRB in August 2020, reference 2020-0803.

The authors have no disclaimers.

The authors report no previous presentations of this work.

