## Supplemental_Appendix_2_FourCorners_FormB for "“We had to put ourselves in their shoes”: Experiences of Medical Students and ObGyn Residents with a Values Clarification Workshop on Abortion"

### Four Corners - Form B

This form assesses the strength which you agree or disagree with a variety of statements about abortion for yourself or someone close to you.

Instructions - Please read the following statements and mark the answers that best reflect your personal beliefs. Please be honest and DO NOT write your name on this sheet. If you are not capable of pregnancy, please respond as though you were a person capable of pregnancy in this activity.

| | Strongly Agree | Agree | Disagree | Strongly Disagree |
|---|---|---|---|---|
| 1. Abortion services should be available to me if I want them. | ○ | ○ | ○ | ○ |
| 2. If I had an abortion, I would be ending a life. | ○ | ○ | ○ | ○ |
| 3. I should be able to have an abortion even if my spouse/ partner wants me to continue the pregnancy. | ○ | ○ | ○ | ○ |
| 4. Liberal abortion laws will lead to me behaving in a more sexually irresponsible way. | ○ | ○ | ○ | ○ |
| 5. If I were young and unmarried, I should be allowed to have an abortion if I wanted one. | ○ | ○ | ○ | ○ |
| 6. If I were a clinician specializing in Ob/Gyn, I would have a responsibility to perform abortions. | ○ | ○ | ○ | ○ |
| 7. If I were a minor, I should be required to get my parents' consent in order to have an abortion. | ○ | ○ | ○ | ○ |
| 8. If I were pregnant and had a terminal disease, I should be counseled to terminate the pregnancy, even if it is desired. | ○ | ○ | ○ | ○ |
| 9. I would not seriously consider the consequences before having an abortion. | ○ | ○ | ○ | ○ |
| 10. I should be able to have a second-trimester abortion (13-24 weeks) if I need one. | ○ | ○ | ○ | ○ |
| 11. If I had an abortion in the second trimester (13-24 weeks), it would be because I was being indecisive. | ○ | ○ | ○ | ○ |
| 12. If I had multiple abortions, I should be encouraged to undergo sterilization. | ○ | ○ | ○ | ○ |
