## Supplemental_Appendix_1_FourCorners_FormA for "“We had to put ourselves in their shoes”: Experiences of Medical Students and ObGyn Residents with a Values Clarification Workshop on Abortion"

### Four Corners - Form A

This form assesses the strength with which you agree or disagree with a variety of statements about abortion for patients in general.

Instructions - Please read the following statements and mark the answers that best reflect YOUR PERSONAL BELIEFS. Please be honest. DO NOT write your name on this sheet.

| | Strongly Agree | Agree | Disagree | Strongly Disagree |
|---|---|---|---|---|
| 1. Abortion services should be available to every person who wants them. | ○ | ○ | ○ | ○ |
| 2. People who have an abortion are ending a life. | ○ | ○ | ○ | ○ |
| 3. A person should be able to have an abortion even if their partner/spouse wants them to continue the pregnancy. | ○ | ○ | ○ | ○ |
| 4. Liberal abortion laws lead to more irresponsible sexual behavior. | ○ | ○ | ○ | ○ |
| 5. Young unmarried people should be allowed to have an abortion if they want one. | ○ | ○ | ○ | ○ |
| 6. Clinicians who specialize in Ob/Gyn have a responsibility to perform abortions. | ○ | ○ | ○ | ○ |
| 7. Minors should be required to get their parents' consent in order to have an abortion. | ○ | ○ | ○ | ○ |
| 8. A pregnant person who has a terminal disease should be counseled to terminate the pregnancy, even if it is desired. | ○ | ○ | ○ | ○ |
| 9. Most people do not seriously consider the consequences before having an abortion. | ○ | ○ | ○ | ○ |
| 10. People should be able to have a second trimester (13-24 weeks) abortion, if they need one. | ○ | ○ | ○ | ○ |
| 11. People who have second-trimester (13-24 weeks) abortions are indecisive. | ○ | ○ | ○ | ○ |
| 12. People who have multiple abortions should be encouraged to undergo sterilization. | ○ | ○ | ○ | ○ |
