## Supplemental_Appendix_3_FINAL_VCAT_InterviewGuide for "“We had to put ourselves in their shoes”: Experiences of Medical Students and ObGyn Residents with a Values Clarification Workshop on Abortion"

**Students**

- Have you read over the information sheet for this research study and do you consent to being a part of this research?
  - We are recording this to transcribe for our research, but we are also using videos of interviews for grant applications and conference submissions, do you consent to having your recording used for these purposes? You may opt out of this and still participate in the research study.
- What medical school do you attend?
- Before the [date of session at that school] values clarification and transformation workshop session on abortion, had you ever participated in the values clarification and transformation workshop?
- Can you describe in a couple of sentences what the workshop was like for you?
- What was it like to hear your beliefs represented by others?
- What did you like about the session?
- What needs improvement?
- What did you like about using a virtual platform for this type of workshop?
- What didn’t you like about the virtual platform?
- Did you find yourself thinking differently about any of your beliefs after the workshop?
  - Do you remember which ones?
  - Was there a rationale that prompted you to think differently?
- Prior to the workshop, what was your general impression of the attitudes and beliefs about abortion held by the other students in your program?
  - Did this change after the workshop? If so, how?
- After participating in the workshop, how do you think your beliefs and your classmates’ beliefs about abortion affect societal stigma or acceptance of abortion?
- During the workshop, did you learn anything that surprised you about your classmates?
- What are *your* views on abortion?
  - Did your views change in any way as a result of the workshop?
- Where there any questions or discussions that made you uncomfortable?
  - Do you remember which one(s)?
- Will your experience in this workshop impact how you care for or counsel patients? If so, how?
- Any other comments or thoughts you’d like to share?

**Residents**

- Have you read over the information sheet for this research study and do you consent to being a part of this research?
  - We are recording this to transcribe for our research, but we are also using videos of interviews for grant applications and conference submissions, do you consent to having your recording used for these purposes? You may opt out of this and still participate in the research study.
- What residency program are you in?
- What year in residency are you?
- Before the [date of session at that institution] values clarification and transformation session on abortion, had you ever participated in the values clarification and transformation workshop?
- Can you describe in a couple of sentences what the workshop was like for you?
- What was it like to hear your beliefs represented by others?
- What did you like about the session?
- What needs improvement?
- What did you like about using a virtual platform for this type of workshop?
- What didn’t you like about the virtual platform?
- Did you find yourself thinking differently about any of your beliefs after the workshop?
  - Do you remember which ones?
  - Was there a rationale that prompted you to think differently?
- Prior to the workshop, what was your general impression of the attitudes and beliefs about abortion held by the other residents in your program?
  - Did this change after the workshop? If so, how?
- After participating in the workshop, how do you think your beliefs and your co-residents’ beliefs about abortion affect societal stigma or acceptance of abortion?
- During the workshop, did you learn anything that surprised you about your co-residents?
- What are *your* views on abortion?
  - Did your views change in any way as a result of the workshop?
- Where there any questions or discussions that made you uncomfortable?
  - Do you remember which one(s)?
- Will your experience in this workshop impact how you care for or counsel patients? If so, how?
- Any other comments or thoughts you’d like to share?

**Facilitator**

- Have you read over the information sheet for this research study and do you consent to being a part of this research?
  - We are recording this to transcribe for our research, but we are also using videos of interviews for grant applications and conference submissions, do you consent to having your recording used for these purposes? You may opt out of this and still participate in the research study.
- What institution do you work for?
- How long have you been in practice/out of residency?
- How long have you been a faculty at this institution?
- Can you tell me how you became involved with this project?
- Prior to your most recent experience facilitating a values clarification and transformation, or VCAT workshop, have you participated in or facilitated a VCAT workshop?
  - If yes, can you briefly share a little bit about that experience?
  - How did that prior experience compare to the current VCAT session?
- Have you led the Four Corners workshop with students, residents, or both?
- The start of this project overlapped with the COVID-19 outbreak and subsequent pandemic. Can you tell me how this impacted your experience developing and delivering the workshop?
- What did you think about using a virtual platform for this workshop?
  - What were some advantages?
  - Some disadvantages?
  - If you were advising someone else designing a similar values clarification project, would you suggest they have it virtually or in person? Why?
- Prior to the workshop, what was your general impression about the attitudes and beliefs about abortion held by the students at your institution?
  - (AND if they ALSO ran session with residents-the residents in your program?) Did this change after the workshop? If so, how?
- Where there any questions or discussions that made you uncomfortable?
  - Do you remember which one(s)?
- After participating in the workshop, how do you think your beliefs and those of your students and residents about abortion affect societal stigma or acceptance of abortion?
- What are *your* views on abortion?
  - Did your views change in any way as a result of the workshop?
- Any other comments/thoughts you’d like to share?
